# Harnessing generative AI to annotate the severity of all phenotypic abnormalities within the Human Phenotype Ontology

**DOI:** 10.1101/2024.06.10.24308475

**Authors:** Kitty B Murphy, Brian M Schilder, Nathan G Skene

**Author notes:** Corresponding author: Kitty B Murphy, Corresponding author: Nathan G Skene.

## Abstract

There are thousands of human phenotypes which are linked to genetic variation. These range from the benign (white eyelashes) to the deadly (respiratory failure). The Human Phenotype Ontology has categorised all human phenotypic variation into a unified framework that defines the relationships between them (e.g. missing arms and missing legs are both abnormalities of the limb). This has made it possible to perform phenome-wide analyses, e.g. to prioritise which make the best candidates for gene therapies. However, there is currently limited metadata describing the clinical characteristics / severity of these phenotypes. With >17500 phenotypic abnormalities across >8600 rare diseases, manual curation of such phenotypic annotations by experts would be exceedingly labour-intensive and time-consuming. Leveraging advances in artificial intelligence, we employed the OpenAI GPT-4 large language model (LLM) to systematically annotate the severity of all phenotypic abnormalities in the HPO. Phenotypic severity was defined using a set of clinical characteristics and their frequency of occurrence. First, we benchmarked the generative LLM clinical characteristic annotations against ground-truth labels within the HPO (e.g. phenotypes in the ‘Cancer’ HPO branch were annotated as causing cancer by GPT-4). True positive recall rates across different clinical characteristics ranged from 89-100% (mean=96%), clearly demonstrating the ability of GPT-4 to automate the curation process with a high degree of fidelity. Using a novel approach, we developed a severity scoring system that incorporates both the nature of the clinical characteristic and the frequency of its occurrence. These clinical characteristic severity metrics will enable efforts to systematically prioritise which human phenotypes are most detrimental to human health, and best targets for therapeutic intervention.

## 0.2 Introduction

Ontologies provide a common language with which to communicate concepts. In medicine, ontologies for phenotypic abnormalities are invaluable for defining, diagnosing, prognosing, and treating human disease. Since 2008, the Human Phenotype Ontology (HPO) has been instrumental in healthcare and biomedical research by providing a framework for comprehensively describing human phenotypes and the relationships between them (Gargano et al., 2024; Köhler et al., 2021). By expanding its depth and breadth over time, the HPO now contains >17500 phenotypic abnormalities across >8600 diseases. Some HPO phenotypes also contain metadata annotations such typical age of onset, frequency, triggers, time course, mortality rate and typical severity. Describing the severity-related attributes of a disease is crucial for both research and clinical care of individuals with rare diseases. When researchers or clinicians are presented with phenotypes that fall outside of their expertise, resources to quickly and reliably retrieve summaries with additional relevant information about these phenotypes are essential. In the clinic, this can help in reaching a differential diagnosis or prioritising the treatment of some phenotypes over others. In research, this information is useful for prioritising targets for causal disease mechanisms, performing large-scale analyses of phenotypic data, and guiding funding agencies when assessing the potential impact and need for research in a given disease area. To date, the HPO has largely relied on manual curation by domain experts. While this approach can improve annotation quality and accuracy, it is both time-consuming and labour-intensive. As a result, less than 1% of terms within the HPO contain metadata such as time course and severity.

Artificial intelligence (AI) capabilities have advanced considerably in recent years, presenting new opportunities to integrate natural language processing technologies into assisting in the curation process. Specifically, there have recently been considerable advances in large language model (LLM) and their application to biomedical problems, in some cases performing as well or better than human clinicians on standardised medical exams and patient diagnosis tasks (Bolton et al., 2024; Cheng et al., 2023; Gu et al., 2021; Labrak et al., 2024; Luo et al., 2022; McDuff et al., 2023; O’Neil et al., 2024; Shin et al., 2020; Singhal, Azizi, et al., 2023, 2023; Singhal, Tu, et al., 2023; Van Veen et al., 2024; Zhang et al., 2023). Recent work has demonstrated that the Generative Pre-trained Transformer 4 (GPT-4) foundation model (OpenAI et al., 2024), when combined with strategic prompt engineering, can outperform even specialist LLMs that are explicitly fine-tuned for biomedical tasks (Nori et al., 2023). In a landmark achievement, GPT-4 was the first LLM to surpass a score of 90% in the United States Medical Licensing Examination (USML) (Nori et al., 2023).

Here, we have used GPT-4 to systematically annotate the severity of 17502 / 17548 (99.7%) phenotypic abnormalities within the HPO. Our severity annotation framework was adapted from previously defined criteria developed through consultation with clinicians (Lazarin et al., 2014). The authors consulted 192 healthcare professionals for their opinions on the relative severity of various clinical characteristics: they used this to create a system for categorising the severity of diseases. Briefly, each healthcare professional was sent a survey asking them to first rate how important a disease characteristic was for determining disease severity, and then to rate the severity of a set of given disease. Using the responses, the authors were able to categorise clinical characteristics into 4 ‘severity tiers’. While characteristics such as shortened lifespan in infancy and intellectual disability were identified as highly severe and placed into tier 1, sensory impairment and reduced lifespan were categorised as less severe and placed into tier 4. Standardised metrics of severity allow clinicians to quickly assess the urgency of treating a given phenotype, as well as prognosing what outcomes might be expected.

To evaluate the consistency of responses generated by GPT-4 793 phenotypes were annotated multiple times. For a subset of phenotypes with known expected clinical characteristics, true positive rates were calculated to assess recall. Additionally, based on the clinical characteristics and their occurrence, we have quantified the severity of each phenotype, providing an example of how these clinical characteristic annotations can be used to guide prioritisation of gene therapy trials. Ultimately, we hope that our resource will be of utility to those working in rare diseases, as well as the wider healthcare community.

## 0.3 Results

### 0.3.1 Annotating the HPO using GPT-4

We employed the OpenAI GPT-4 model with Python to annotate 17502 terms within the HPO (v2024-02-08) (Gargano et al., 2024; Köhler et al., 2021). Our annotation framework was developed based on previously defined criteria for classifying disease severity (Lazarin et al., 2014). We sought to evaluate the impact of phenotypes on factors including intellectual disability, death, impaired mobility, physical malformations, blindness, sensory impairments, immunodeficiency, cancer, reduced fertility, and congenital onset. Through prompt design we found that the performance of GPT-4 improved when we incorporated a scale associated with each clinical characteristic and required a justification for each response. For each clinical characteristic, we asked about the frequency of its occurrence - whether it never, rarely, often, or always occurred. Framing the queries in this way served two purposes. First, this helped to constrain the responses of GPT-4 to a specific range of values, making answers more consistent and amenable to downstream data analysis. Second, it served to overcome one of the main limitations noted by Lazarin et al. (2014) as they did not collect information on how the frequency of each disease affected their decision making when generating severity annotations.

Clinical characteristic occurrence varied across annotation categories. >50% of phenotypes never caused blindness, sensory impairments, immunodeficiency, cancer, reduced fertility or intellectual disability. Only a minority of phenotypes (21.7%) never had a congenital onset, which is expected as rare disorders tend to be early onset genetic conditions (Fig. 1).

**Figure 1:**
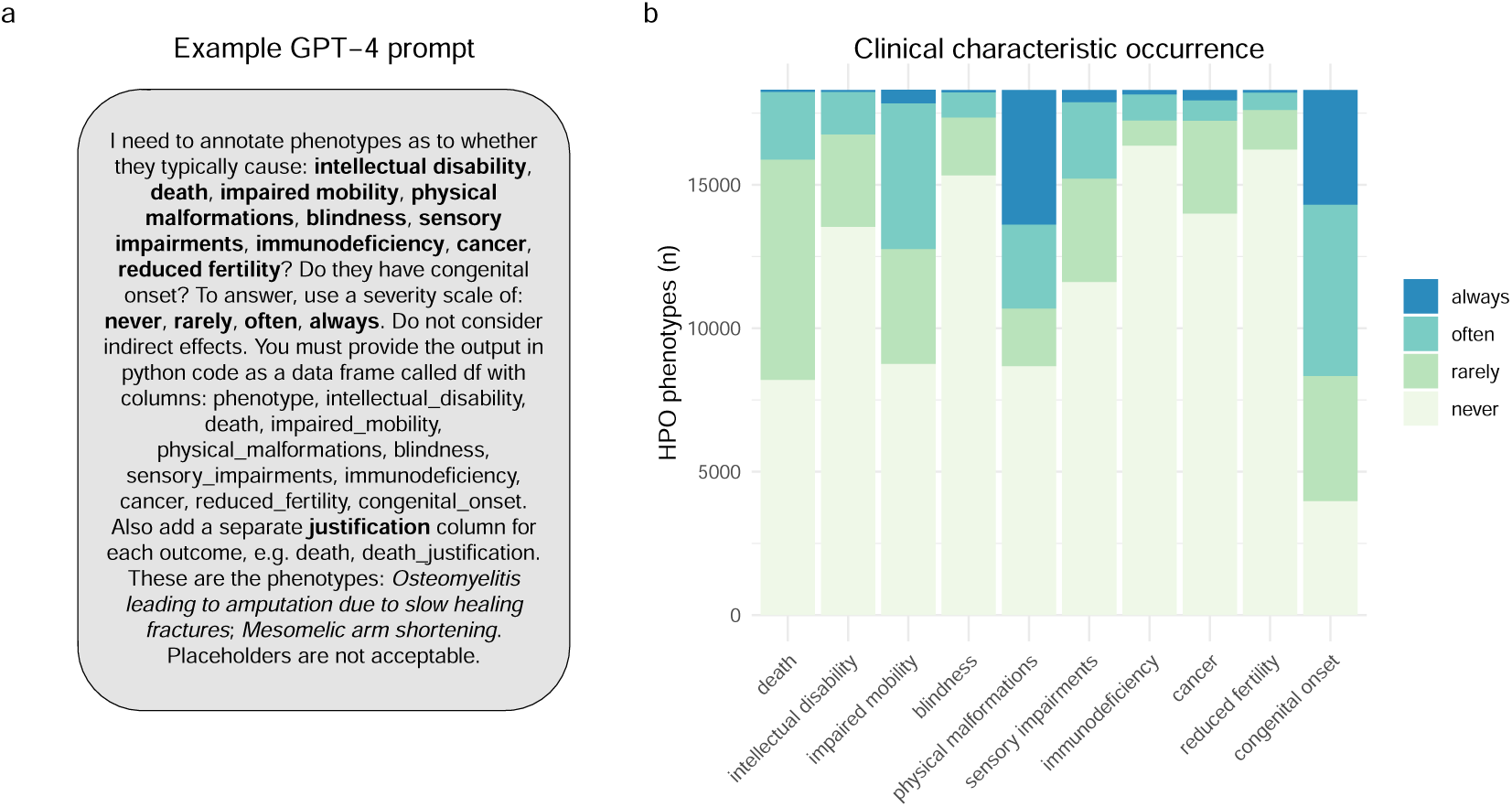
GPT-4 was able to annotate all human phenotypes based on whether they are always/often/rarely/never associated with different clinical characteristics. **a** An example of the prompt input given to to GPT-4. The phenotypes listed in the second to last sentence (*italicised*) were changed to allow all HPO phenotypes to be annotated. **b** Stacked bar plot showing the proportion of the occurrence of each clinical characteristic across all annotated HPO phenotypes. The terms shown on the x-axis are the clinical characteristics for which GPT-4 was asked to determine whether each phenotype caused them.

**Figure 2:**
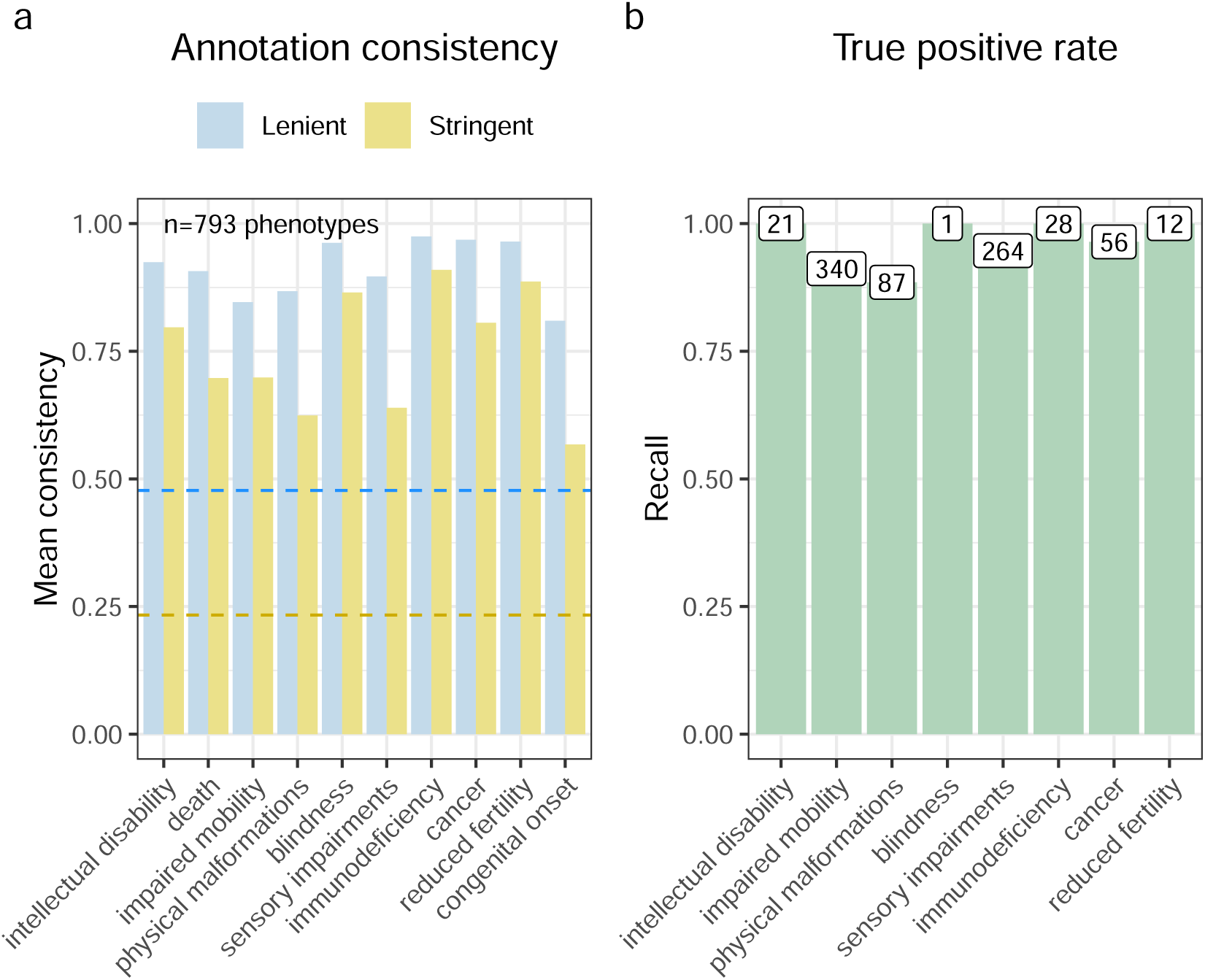
GPT-4 annotations are consistent and accurate across annotations. **a** Barplot showing the annotation consistency within phenotypes that were annotated more than once. In the lenient metric, annotations were collapsed into two groups (‘always’/‘often’ and ‘never’/‘rarely’). For a given clinical characteristic within a given phenotype, if an annotation was always within the same group it was considered consistent. In the stringent metric, all four annotation categories were considered to be different from one another. Thus, annotations were only defined as consistent if they were all identical. The blue dashed line indicates the probability of two annotations being consistent by chance in the lenient metric (∼1/2). The gold dashed line is the probability of two annotations being consistent by chance in the stringent metric (∼1/4). **b** Bar plot of the true positive rate for each annotation. The labels above each bar indicate the number of phenotypes tested.

Less than 1% of phenotypes always directly resulted in death (n=71), such as ‘Stillbirth’, ‘Anencephaly’ and ‘Bilateral lung agenesis’. Meanwhile, 9707 phenotypes were annotated as often or rarely causing death. 7880 phenotypes were annotated as never causing death. Examples of phenotypes that never cause death included 34 unique forms of syndactyly, a non-lethal condition that causes fused or webbed fingers (occurring 1 in 1,200–15,000 live births). While not life-threatening itself, syndactyly is a symptom of genetic disorders that can cause life-threatening cardiovascular and neurodevelopmental defects, such as Apert Syndrome (Garagnani & Smith, 2013). This example highlights the ability of GPT-4 to successfully distinguish between phenotypes that directly cause lethality, and those that are often associated with diseases that cause lethality.

#### 0.3.2 Annotation consistency and recall

To assess annotation consistency, we queried GPT-4 with a subset of the HPO phenotypes multiple times (n=793 unique phenotypes). We employed two different metrics to determine the *consistency rate*. The first, less stringent metric, defined consistency as the duplicate annotations being either ‘always’ and ‘often’, or ‘never’ and ‘rarely’. The second, more stringent metric, required exact agreement in annotation occurrences, e.g. ‘always’ and ‘always’. For the less stringent metric, duplicated phenotypes were annotated consistently at a rate of at least 80%, and for the more stringent metric, the lowest consistency rate was 57% for congenital onset. An example of where annotations were inconsistent was for the HPO term ‘Acute leukaemia’. One time, GPT-4 annotated it as often causing impaired mobility, giving the justification that ‘weakness and fatigue from leukaemia and its treatment can impair mobility’. The other time, GPT-4 annotated it as rarely causing impaired mobility, giving the justification that ‘acute leukaemia rarely impairs mobility directly’. Despite specifying in the prompt for GPT-4 not to take into consideration indirect effects, this is an example of where it failed to do so.

We also reasoned that GPT-4 would be better able to give consistent answers for more specific phenotypes lower in the ontology, as they are more likely to have a single cause. We found that the stringent consistency rate did indeed significantly improve with greater HPO ontology depth (*X_Pearson_*^2^ =22.17, *V̂_Cramer_*=0.03, p=0.05). See Figure 5 for a visual representation of this relationship.

In order to evaluate the validity of the annotations, we calculated a true positive rate. This involved identifying specific branches within the HPO that would contain phenotypes that would reliably indicate the presence of certain conditions. For instance, the phenotypes ‘Decreased fertility in females’ and ‘Decreased fertility in males’ should often or always cause reduced fertility. We observed an encouraging true positive rate exceeding 88% across in every clinical characteristic and achieving perfect recall (100%) in 4/8 characteristics.

The lowest true positive rate was observed for physical malformations, with 88.5% recall across 87 HPO phenotypes. Some cases in which the GPT-4 annotations disagreed with the HPO ground truth included: ‘Angioma serpentinum’, ‘Nevus anemicus’, ‘Pulmonary arteriovenous fistulas’. In the case of ‘Angioma serpentinum’ it provided the justification that ‘No known association with physical malformations’. In another instance, GPT-4 noted that ‘Nevus anemicus’ is ‘Limited to hypopigmented skin patch; no other malformations.’. This indicates that while technically incorrect according to our predefined benchmarks, a case could in fact be made that mild skin conditions do not rise to the level of physical malformations.

This high level of recall underscores the robustness of our annotations and the reliability of the HPO framework in capturing clinically relevant phenotypic information. However, we acknowledge that the number of testable true positive phenotypes for some of these categories are low, especially ‘blindness’ for which there is only 1 phenotype in the HPO (after excluding terms pertaining to colour or night blindness). Furthermore, some of the true positive phenotypes are lexically similar to the name of the clinical characteristic itself. In these cases, annotating ‘Severe intellectual disability’ as always causing intellectual disability is a relatively trivial task. Nevertheless, even these scenarios provide a clear and interpretable benchmark for the model’s performance. In addition, were numerous phenotypes with lexically non-obvious relationships to the clinical characteristic that were annotated correctly by GPT-4. For example, ‘Molar tooth sign on MRI’ (a neurodevelopmental pathology observed in radiological scans) was correctly annotated as causing intellectual disability.

#### 0.3.3 Quantifying phenotypic severity

While individual annotations are informative, we wanted to be able to distil the severity of each phenotype into a single score. Quantifying the overall severity of phenotypes can have important implications for diagnosis, prognosis, and treatment. It may also guide the prioritisation of gene therapy trials for phenotypes with the most severe clinical characteristics and thus the most urgent need. Importantly, the values reflected the severity of each clinical characteristic based on both the type of characteristic itself and its frequency within a particular phenotype. For instance, a phenotype always causing death would have a higher multiplied value than a phenotype often causing reduced fertility (see Table 2). First, we created a dictionary to map each clinical characteristic (e.g. blindness) and its frequency (always, often, rarely, never) to numeric values from 0-3. Then, the clinical characteristic values were multiplied by weights. Next, we computed an average score for each phenotype by aggregating the multiplied values across all clinical characteristics and then calculating the mean. This was then normalised by the theoretical maximum severity score, so that all phenotypes were on a 0-100 severity scale (where 100 is the most severe phenotype possible). This average normalised score represents the overall severity of the phenotype based on the severity of its individual clinical characteristics.

Based on these scores we evaluated the top 50 severe phenotypes. One of the most severe phenotype was ‘Anencephaly’ (HP:0002323) with a composite severity score of 45. Anencephaly is a birth defect where the baby is born without a portion of its brain and skull, often these babies are stillborn. In fact, many of the most severe phenotypes were related to developmental brain and neural tube defects. Comparison of the severity scores for each response, across the clinical characteristics annotated, revealed consistent trends: as the response of the clinical characteristic increased (from never to always), the severity score also increased (Supplementary Fig. 7). We also evaluated the severity score distribution by HPO branch and calculated the mean severity score using all phenotypes within each major HPO branch (Fig. 6). The HPO branch with the greatest mean severity score was ‘Abnormal cellular phenotype’ (mean=17), followed by ‘Neoplasm’ (mean=16.7), which would include the highly ranked phenotypes seen in Figure 3.

**Figure 3:**
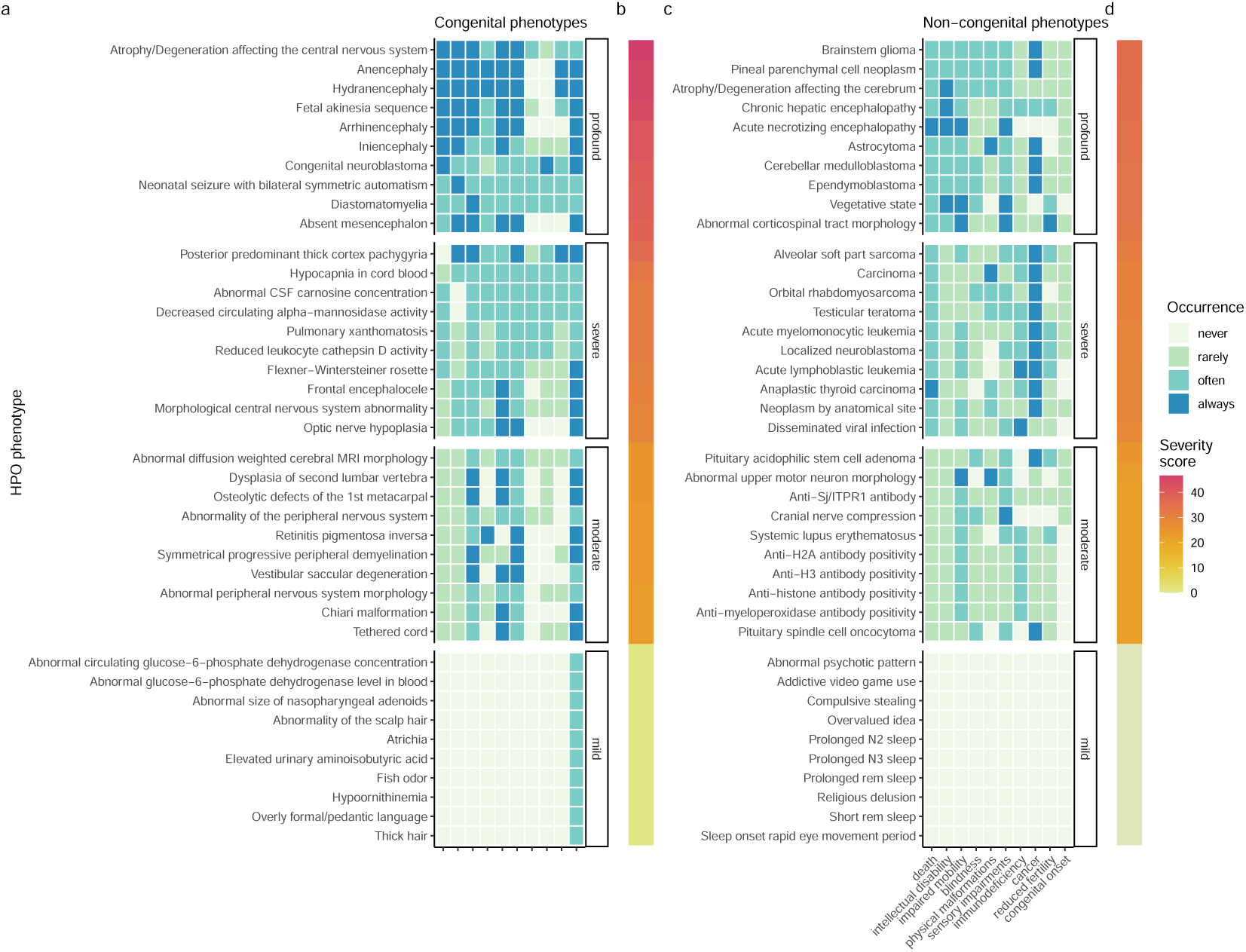
Quantifying the severity of HPO phenotype annotations highlights the most impactful conditions. Heatmap of 10 represetantive phenotypes from each severity class (Profound, Severe, Moderate, Mild) stratified by whether the phenotypes are often/always congenital (**a**-**b**) or rarely/never congenital (**c**-**d**). Continuous severity scores are shown as bars (**b**,**d**) and were calculated by multiplying the numeric values assigned to each clinical characteristic according to Table 2. The average normalised score, representing overall phenotype severity on a 0-100 scale, was calculated by aggregating the multiplied values and normalising by the theoretical maximum severity score. The x-axes show each of the clinical characteristics. All data for this figure, as well as justifications for each annotation, can be found in Table 3.

#### 0.3.4 Severity classes

While the continuous severity score is a helpful metric, there may be some use cases where a categorical classification of severity is more immediately useful. In work by Lazarin et al. (2014), the authors defined severity classed using a simple decision tree based on the individual severity annotations. We approximated this approach using our GPT-4 annotations. This categorical approach showed a strong degree of positive correspondence with the continuous severity score (*ŵp*^2^=0.88, p<2.2e-308). In other words, severity score increased with severity class level (mild < moderate < severe < profound) as expected. The distribution of severity classes is shown in Figure 9.

#### 0.3.5 Correlations between clinical characteristic severity metrics

We found that some clinical characteristic severity metrics were correlated with one another, with a mean Pearson correlation of 0.2 across all individual metrics (see Figure 8). In particular, blindness and sensory impairment were highly correlated with one another (r=0.62, p=0). Some metrics drove the composite severity score more than other, which is a reflection of both our per-metric weighting scheme, response type frequencies, and the correlation structure between metrics. Overall, impaired mobility seemed to be the strongest driver of the composite severity score with a Pearson correlation of 0.6001824, followed by intellectual disability (r=0.59) and death (r=0.56).

#### 0.3.6 Congenital onset by HPO branch

Next, we assessed the distribution of congenital onset across HPO branches (Fig. 4). We found that the Abnormality of prenatal development or birth branch contained the greatest proportion of phenotypes that were always congenital (70.15%), followed by Abnormality of the musculoskeletal system (45.34%) and Growth abnormality (37.62%). This is concordant with the expectation that these phenotypes should largely be congenital. The HPO branches with the least commonly congenital phenotypes were Constitutional symptom (0%), Abnormality of the thoracic cavity (0%), and Phenotypic abnormality (0%). ‘Constitutional symptom’ is a fairly broad term defined as *‘A symptom or manifestation indicating a systemic or general effect of a disease and that may affect the general well-being or status of an individual.’* Examples include ‘Fatigue’ ‘Exercise intolerance’, ‘Hot flashes’ and ‘Sneeze’.

**Figure 4:**
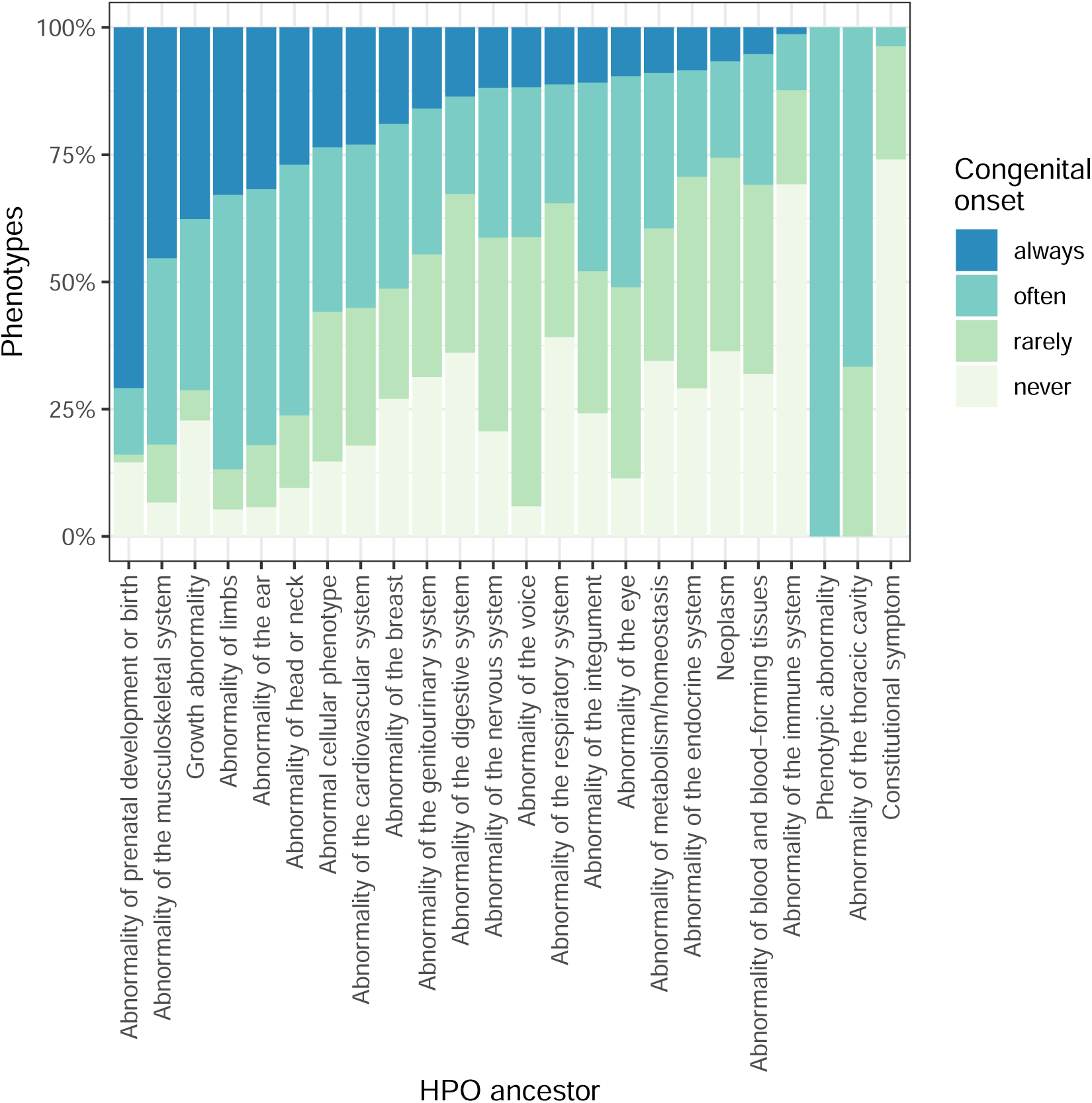
Distribution of congenital onset across HPO branches. The y-axis shows the proportion of phenotypes that are always/often/rarely/never congenital. The x-axis shows the HPO branch, orderered from highest to lowest proportion of always congenital phenotypes.

## 0.4 Discussion

Phenotype severity annotations have utility across a wide variety of applications in both the clinic and research. In clinical settings, severity annotations can be used to prioritise the treatment of some phenotypes over others in patients with complex presentations, avoid administering contraindicated drugs, and prognosing potential health outcomes. In research settings, severity annotations can be used to identify phenotypes that have a large impact on patient outcomes and yet are currently understudied. They may also be used to help design new experiments and studies, or even provide new insights into the underlying aetiology of the disease by making expert-level summaries more immediately accessible to the wider research community.

The creation and annotation of biomedical knowledge has traditionally relied on manual or semi-manual curation by human experts (Gargano et al., 2024; Köhler et al., 2021; Mungall et al., 2017; Ochoa et al., 2021; Putman et al., 2024). Performing such manual curation and review tasks at scale is often infeasible for human biomedical experts given limited time and resources. LLMs have the capacity to effectively encode, retrieve, and synthesise vast amounts of diverse information in a highly scalable manner (OpenAI et al., 2024; Singhal, Azizi, et al., 2023; Van Veen et al., 2024). This makes them powerful tools that can be applied in a rapidly expanding variety of scenarios, including medical practice, research and data curation (Caufield et al., 2023; O’Neil et al., 2024; Pan et al., 2023; Singhal, Azizi, et al., 2023; Toro et al., 2023).

Here, we introduce a novel framework to leverage the current best-in-class LLM, GPT-4 (OpenAI et al., 2024), to systematically annotate the severity of 17502 phenotypic abnormalities within the HPO. By employing advanced AI capabilities, we have demonstrated the feasibility of automating this process, significantly enhancing efficiency without substantially compromising accuracy. Our validation approach yielded a high true positive rate exceeding 88% across the phenotypes tested. Furthermore, our approach can be readily adapted and scaled to accommodate the growing volume of phenotypic data. In total, the entire study cost $296.27 in queries to the OpenAI API. While we do not have a direct comparison, this likely represents a extremely small fraction of the total costs of such a study if performed manually by human experts charging at an hourly rate. Even if all human annotations were provided on a volunteer basis, this would still require hundreds if not thousands of hours of cumulative manual human labour. Using our approach, severity annotations for the entire HPO can be generated automatically at a rate of ∼100 phenotypes/hour. Further optimisation of the annotation process and increased API rate limits could potentially accelerate this even further.

Throughout this study, we observed that GPT-4 was capable of reliably recovering deep semantic relationships from the medical domain, far beyond making superficial inferences based on lexical similarities. An excellent example of this is the phenotype ‘Molar tooth sign on MRI’ (HP:0002419; severity score=25.56), which GPT-4 annotated as causing intellectual disability. At first glance, we ourselves assumed this was a false positive as the term appeared to be related to dentition. However, upon further inspection we realised that molar tooth sign is in fact a pattern of abnormal brain morphology that happens to bear some resemblance to molar dentition when observed in radiological scans. This phenotype is a known sign of neurodevelopmental defects that can indeed cause severe intellectual disability (Gleeson et al., 2004).

In addition to rapidly synthesising and summarising vast amounts of information, LLMs can also be steered to provide justifications for each particular response. This makes LLMs amenable to direct interrogation as a means of recovering explainability, especially when designed to retain information about previous requests and interactions as they use these to iteratively improve and update their predictions (Janik, 2024). This represents a categorical advance over traditional natural language processing models based on more shallow forms of statistical or machine learning (e.g. Term Frequency-Inverse Document Frequency (Jones, 1972), Word2vec (Mikolov et al., 2013)) which lack the ability to provide chains of causal reasoning to justify their predictions. This highlights the fundamental trade-off between simpler models with high explainability (the ability humans to understand the inner workings of the model) but low interpretability (the ability of humans to trace the decision process of the model, analogous to human ‘reasoning’), and deeper more complex models with low explainability but high interpretability (Marcinkevičs & Vogt, 2023).

A key contribution of our study is the introduction of a quantitative severity scoring system that integrates both the nature of the clinical characteristic and the frequency of its occurrence. By encoding the concept of severity in this way, we are able to prioritise phenotypes based on their impact on patients. The methodology allowed us to transition from low-throughput qualitative assessments of severity (e.g. Lazarin et al. (2014)) to high-throughput quantitative assessments of severity. One of the most severe phenotypes in the HPO is ‘Fetal akinesia sequence’ (FAS; HP:0001989, severity score= 43.9), and extremely rare condition that is almost always lethal. FAS is a complex, multi-system phenotype that can be caused by at least 24 different genetic disorders. Despite the complex and heterogeneous aetiology of this phenotype, GPT-4 was able to provide accurate annotations alongside explainable justifications for those annotations (see Table 4). For example, this phenotype almost always results in death, either *in utero* or shortly after birth. Not only did GPT-4 correctly provide the annotation death as ‘always’, when asked whether FAS causes sensory impairments it provided the response ‘always’ with the justification ‘Fetal akinesia sequence typically results in severe sensory impairment due to neurodevelopmental disruption.’ Neurodevelopmental disruption is indeed a hallmark component of FAS (e.g. hydrocephalus, cerebellar hypoplasia) that causes severe impairments across multiple sensory systems (Chen, 2012). This demonstrates that GPT-4 was able to recover the correct chain of causality from phenotype to clinical characteristic.

Our findings highlight the potential of this next generation of natural language processing technologies in significantly contributing to the automation and refinement of data curation in biomedical research. These results have a large number of useful real-world applications, such as prioritising gene therapy candidates (Murphy et al., 2023) and guiding clinical decision-making in rare diseases. It may also be used as tool to help inform policy decisions and funding allocation by healthcare or governmental institutions. This of course would need to be in consultation with subject matter medical experts, patients, advocates and biomedical ethicists before reaching a final decision. Nevertheless, access to succinct, interpretable, and semi-quantitative severity annotations may encourage key decision makers with limited time to review individual proposals to pay heed to phenotypes and diseases that would otherwise be overlooked. As the HPO and the broader literature continue to grow over time, our automated AI-based approach can easily be repeated to keep pace with the rapidly evolving biomedical landscape. Furthermore, it can be extended to produce different sets of annotations or be used with any other ontology. Additional use cases include gathering data on the prevalence of each phenotype to approximate their social and financial costs.

One key limitation of our study is the fact that we did not explicitly interrogate GPT-4 to assess how the availability of treatments affected the annotations it produced. For example, there are some very severe conditions for which highly effective treatments and early detection screens are widely available (e.g. syphilis, some forms of melanoma), thus rendering them fully treatable or even curable provided access to modern healthcare. It would therefore be useful to further interrogate GPT-4 to uncover how the availability of treatments influences its responses. Many of our findings here seem to indicate that GPT-4 does take into account quality of care to the extent that health services increase the likelihood of desired outcomes. For example, many of the cancer phenotypes are justified as always or often causing death unless detected and treated early in the disease course. On the other hand, some cancers are justified as rarely causing death if appropriate treatment is provided, which may not always be the case for individuals or populations with access to less access to quality healthcare services. Future efforts could more explicitly ask GPT-4 whether the phenotype would cause death with no or suboptimal treatment.

Another limitation with the present dataset is that phenotypes themselves can manifest with different degrees of severity, in the sense that they are more pronounced or intense. For example, sensitivity to light could range from a mild inconvenience to a severe disability that prevents the individual from leaving their home during the day. The effect of onset (beyond congenital vs. non-congenital) and time course (acute, slowly progression, relapse-remitting) were also not explicitly considered. Finally, we did not ask GPT-4 to consider phenotypes as they present within particular diseases. For example, while the phenotype ‘Hypertension’ may be mild to moderate in the general population and not present until middle-age, it can also present early in life as very severe in the context of a rare genetic disorder such as Liddle syndrome. Future work could explore these nuances in more detail.

In addition to these technical challenges, there are multiple factors that need to be considered when trying to prioritise phenotypes for their suitability for gene therapy development. First, while we have attempted to formalise severity here, this is an inherently subjective concept that may vary considerably across different individuals and contexts. For instance, one could ask whether a condition that always causes death is worse than a condition that causes a lifetime of severe disability (e.g. paralysis, blindness, intellectual disability). Metrics such as quality-adjusted life years (QALYs) have been proposed in the past to address these dilemmas by defining health as a function of both the length and quality of life (Prieto & Sacristán, 2003). With regards to the financial burden of diseases, in some situations phenotypes which require many years of expensive medical care may be prioritised over those that result in extremely early onset lethality and little opportunity for therapeutic intervention. Another factor that affects the viability of a therapeutic program is the speed, cost and other practical considerations of a clinical trial. For instance, measuring risk of ageing-related respiratory failure over a ten-year period may be impractical in some cases. However, testing for total reversal of an existing severe phenotype could potentially yield faster and more immediately impactful results. If performed in close collaboration with medical ethicists, governmental organisations, advocacy groups and patient families, such cost/benefit assessments could be aided by LLMs through the scalable gathering of relevant data. As AI capabilities continue to advance, the range of applications in which they can be used effectively will continue to grow.

While our study demonstrates the feasibility and utility of AI-driven phenotypic annotation, several limitations must be acknowledged. The reliance on computational algorithms may introduce biases or inaccuracies inherent to the training data, necessitating ongoing validation and refinement of our approach. Additionally, our severity scoring system, while comprehensive, may not capture the full spectrum of phenotypic variability or account for complex gene-environment interactions. Future research should focus on further optimising AI-driven annotation methodologies, incorporating additional data modalities such as genomic and clinical data to enhance accuracy.

In conclusion, our study represents a significant step towards harnessing the power of AI to advance phenotypic annotation and severity assessment across all rare diseases. This resource aims to provide researchers and clinicians with actionable insights that can inform rare disease research and improve the lives of individuals affected by rare diseases.

## 0.5 Methods

### 0.5.1 Annotating the HPO using OpenAI GPT-4

We wrote a Python script to iteratively query GPT-4 via the OpenAI application programming interface (API). The ultimately yielded consistently formatted annotations for 17502 terms within the HPO. Our annotation framework was developed based on previously defined criteria for classifying disease severity (Lazarin et al., 2014). We sought to evaluate whether each phenotype directly caused a given severity-related clinical characteristic, including: intellectual disability, death, impaired mobility, physical malformations, blindness, sensory impairments, immunodeficiency, cancer, reduced fertility, and/or had a congenital onset. Through prompt engineering we found that the performance of GPT-4 improved when we incorporated a scale associated with each clinical characteristic and required a justification for each response. We asked how frequently the given phenotype directly causes each clinical characteristic - whether it never, rarely, often, or always occurred. This design helps to constrain the potential responses of GPT-4 and thus make it more amenable to machine-readable post-processing. It also serves to address one of its key limitations from the Lazarin et al. (2014) survey, namely the lack information on how clinical characteristic frequency affected the clinicians’ severity annotations. Here, we can instead use the frequency values to generate more precise annotations and downstream severity ranking scores.

Furthermore, our prompt design revealed that the optimal trade-off between the number of phenotypes and performance (in terms of producing the desired annotations, and adhering to the formatting requirements) was achieved when inputting no more than two or three phenotypes per prompt. An example prompt can be seen in Figure 1. Thus, only two phenotypes were included per prompt in order to 1) avoid exceeding per-query token limits, and 2) prevent the breakdown of GPT-4 performance due to long-form text input, which is presently a known limitation common to many LLMs including GPT-4 (Wei et al., 2024).

### 0.5.2 Calculating the true positive rate

A true positive rate was calculated as a measure of the recall of the GPT-4 annotations. This was achieved by identifying specific branches within the HPO that would contain phenotypes that would reliably indicate the occurrence of certain clinical characteristics, and using all descendants of this HPO branch as true positives. For example, all descendants of the terms ‘Intellectual disability’ (HP:0001249) or ‘Mental deterioration’ (HP:0001268) should be annotated as always or often causing intellectual disability (Table 1).

**Table 1:**
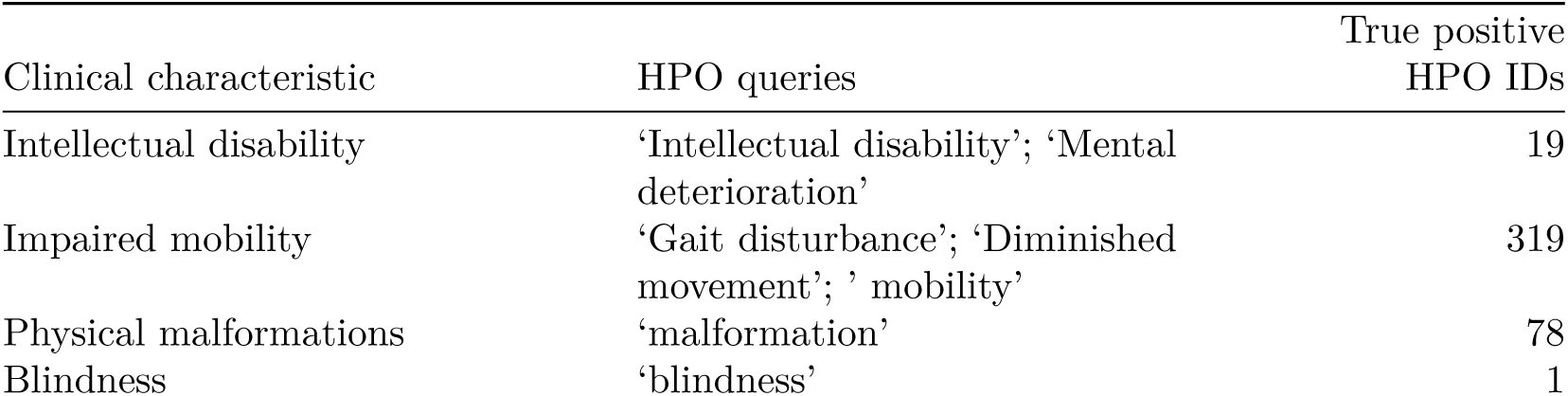
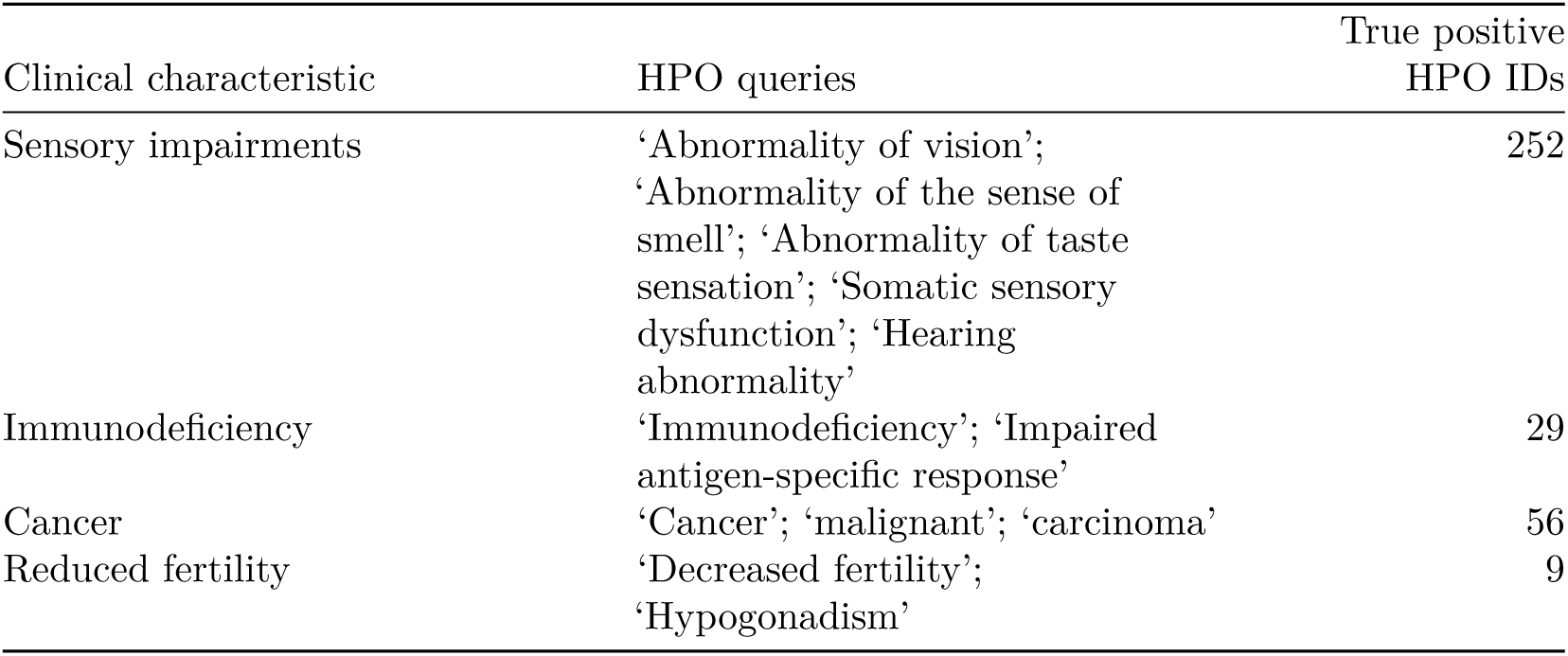
The HPO branches and their descendants used as true positives for each clinical characteristic.

### 0.5.3 Quantifying phenotypic severity

The GPT-4 generated clinical characteristic occurrences were converted into a semi-quantitative scoring system, with ‘always’ corresponding to 3, ‘often’ to 2, ‘rarely’ to 1, and ‘never’ to 0. These scores were then weighted by a severity metric on a scale of 1-5, with 5 representing the highest severity, as determined by the provided clinical characteristics (Table 2). Subsequently, the weighted scores underwent normalisation to yield a final quantitative severity score ranging from 0-100, with 100 signifying the maximum severity score attainable.

**Table 2:** Weighted scores for each clinical characteristic and GPT-4 response category.

| Clinical characteristic | Always (3) | Often (2) | Rarely (1) | Never (0) |
| --- | --- | --- | --- | --- |
| Death (6) | 18 | 12 | 6 | 0 |
| Intellectual disability (5) | 15 | 10 | 5 | 0 |
| Impaired mobility (4) | 12 | 8 | 4 | 0 |
| Blindness (4) | 12 | 8 | 4 | 0 |
| Physical malformations (3) | 9 | 6 | 3 | 0 |
| Sensory impairments (3) | 9 | 6 | 3 | 0 |
| Immunodeficiency (3) | 9 | 6 | 3 | 0 |
| Cancer (3) | 9 | 6 | 3 | 0 |
| Reduced fertility (1) | 3 | 2 | 1 | 0 |
| Congenital onset (1) | 3 | 2 | 1 | 0 |

Let us denote:

- *p*: a phenotype in the HPO.
- *j*: the identity of a given annotation metric (i.e. clinical characteristic, such as ‘intellectual disability’ or ‘congenital onset’).
- *W_j_*: the assigned weight of metric *j*.
- *F_j_*: the maximum possible value for metric *j* (equivalent across all *j*).
- *F_pj_* : the numerically encoded value of annotation metric *j* for phenotype *p*.
- *NSS_p_*: the final composite severity score for phenotype *p* after applying normalisation to align values to a 0-100 scale and ensure equivalent meaning regardless of which other phenotypes are being analysed in addition to *p*. This allows for direct comparability of severity scores across studies with different sets of phenotypes.

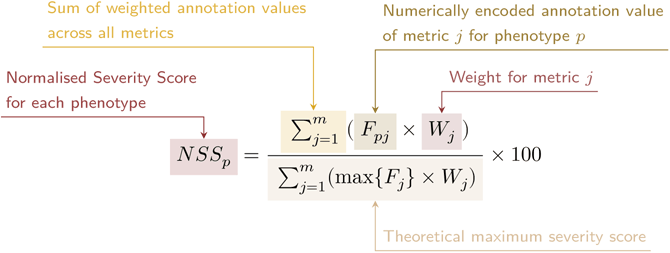

### 0.5.4 Severity classes

The decision tree algorithm used in Lazarin et al. (2014) was adapted here for use with the GPT-4 clinical characteristic annotations. This algorithm first assigned each clinical chacteristic to a tier, where Tier 1 indicated the most severe clinical characteristics and Tier 4 indicated the least severe clinical characteristics (‘death’=1, ‘intellectual disability’=1, ‘impaired mobility’=2, ‘physical malformations’=2, ‘blindness’=3, ‘sensory impairments’=3, ‘immunodeficiency’=3, ‘cancer’=3, ‘reduced fertility’=4). If a phenotype often or always caused more than one Tier 1 clinical characteristic, it was assigned a severity class of “Profound”. If the phenotype often or always caused only one Tier 1 clinical characteristic, it was assigned a severity class of “Severe”. A “Severe” class assignment was also assigned if the phenotype often or always caused three or more Tier 2 and Tier3 clinical characteristics. If the phenotype often or always caused at least one Tier 2 clinical characteristic, it was assigned a severity class of “Moderate”. All remaining phenotypes were was assigned a severity class of “Mild”. In cases where the phenotype mapped to more than one class, only the most severe class was used. This procedure is implemented within the function HPOExplorer::gpt_annot_class.

### 0.5.5 Correlations between clinical characteristic severity metrics

To assess the correlation structure between each clinical characteristic severity metric, as well as between the composite severity score and each metric, we computed Pearson correlation coefficients for all pairwise combinations of these variables using the numerically encoded metric values. The correlation matrix was visualised using a heatmap, with the colour intensity representing the strength of the correlation (Figure 8).

## 0.6 Data and code availability statement

All code and data used in this study are available on GitHub at: https://github.com/neurogenomics/gpt_hpo_annotations

The GPT-4 clinical characteristic annotations for all HPO phenotypes are made available through the R function HPOExplorer::gpt_annot_read or in CSV format at: https://github.com/neurogenomics/gpt_hpo_annotations/tree/master/data

A fully reproducible version of this Quarto manuscript can be found at: https://github.com/neurogenomics/gpt_hpo_annotations/blob/master/manuscript.qmd

## 0.7 Acknowledgements

We would like to thank members of the Monarch Initiative for their insight and feedback throughout this project. In particular, Peter Robinson.

## 0.8 Supplementary Materials

### 0.8.1 Supplementary Figures

**Figure 5:**
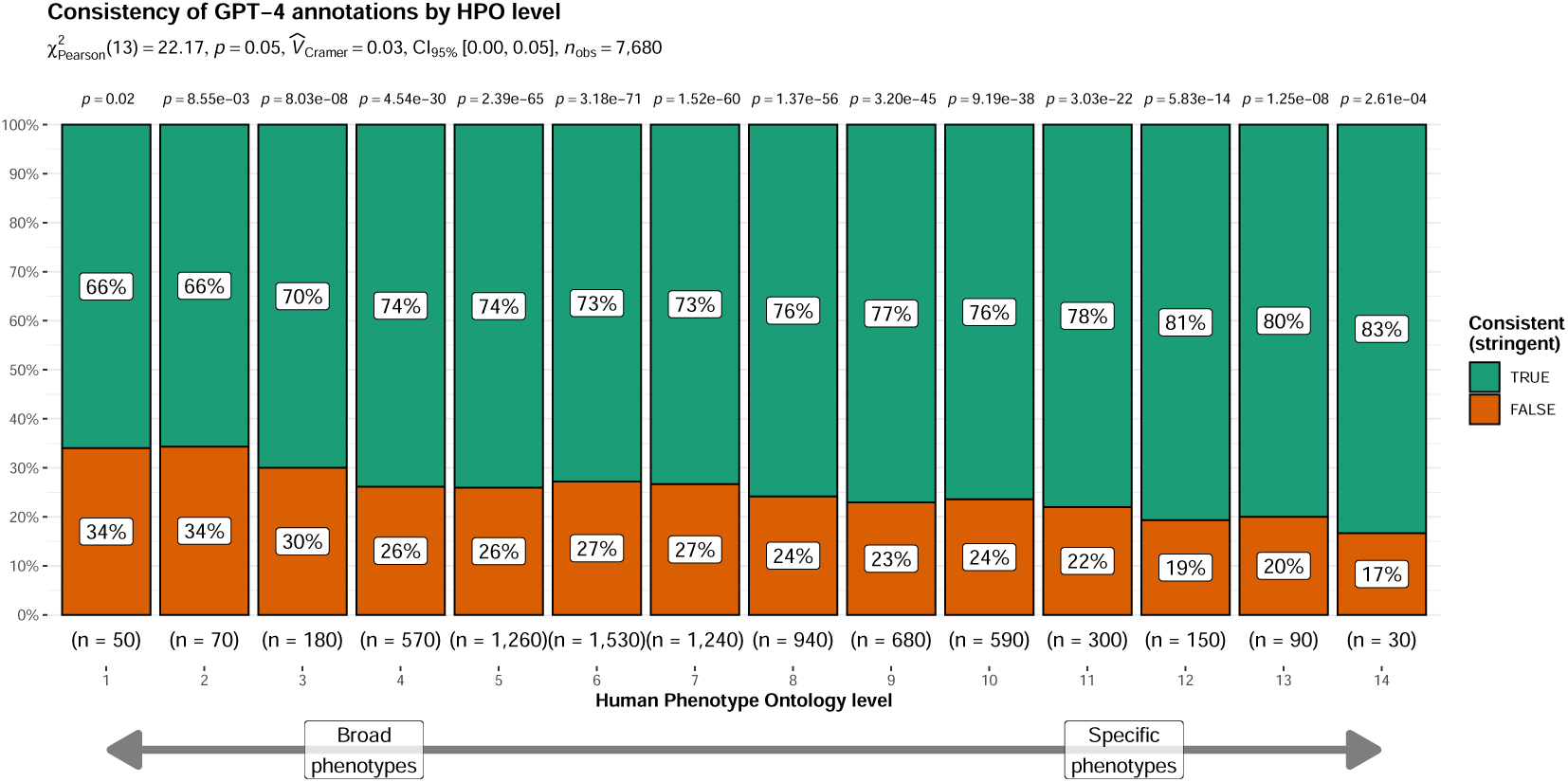
Relationship between the consistency of GPT-4 clinical characterstic annotations (using the stringent criterion) and the level of each phenotype within the HPO ontology (with the number of phenotypes in parentheses). Greater ontology levels (x-axis) indicate more specific phenotypes. The subtitle indicates summary statistics for the overall relationship between HPO level and the proportion of phenotypes that were annotated consistently. The p-values above each bar indicate whether the distribution of consistent/inconsistent annotations, within a given HPO level, significantly deviate from the expected null distribution.

**Figure 6:**
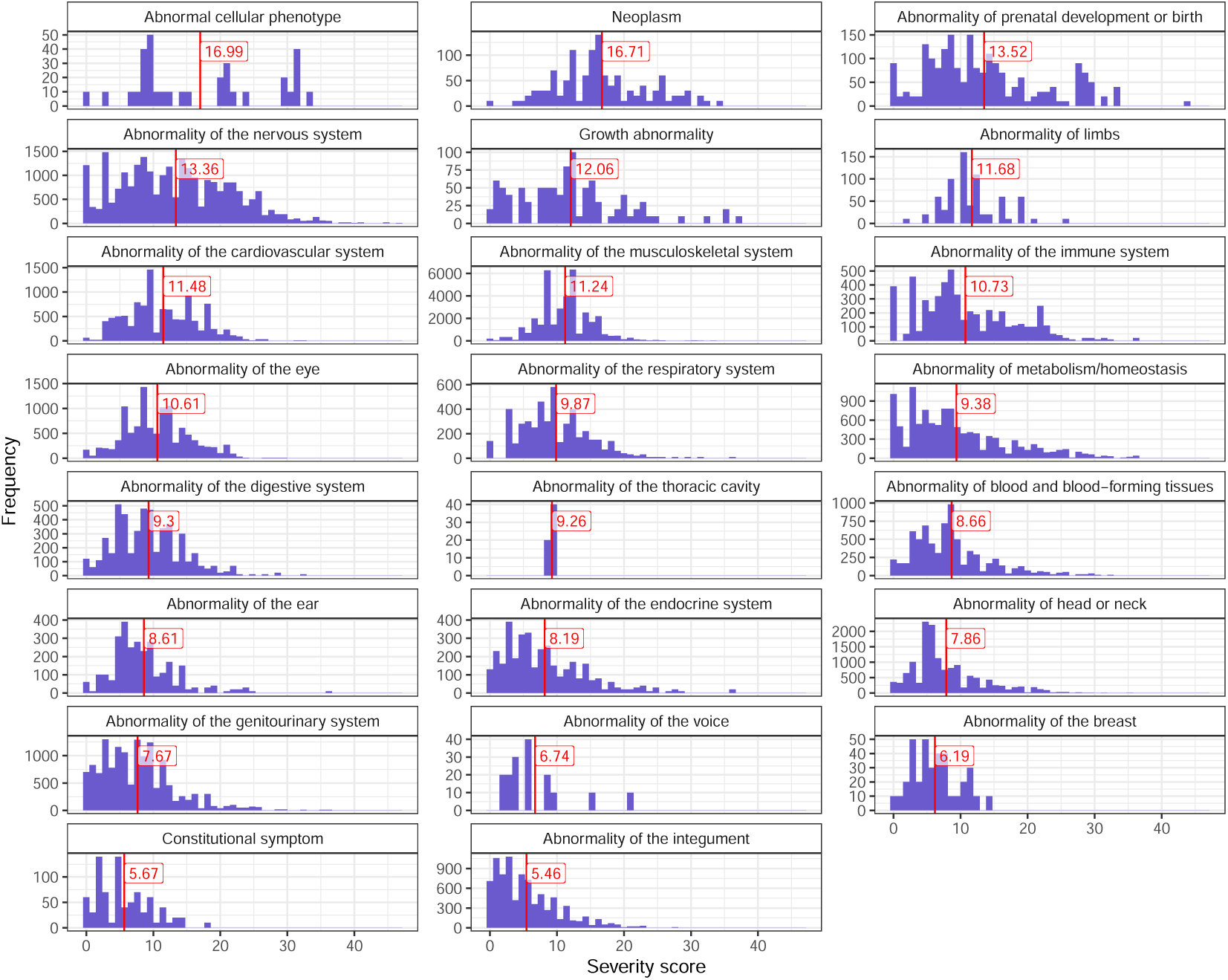
Distribution of the composite GPT-4 severity score of the severity scores for all HPO terms.

**Figure 7:**
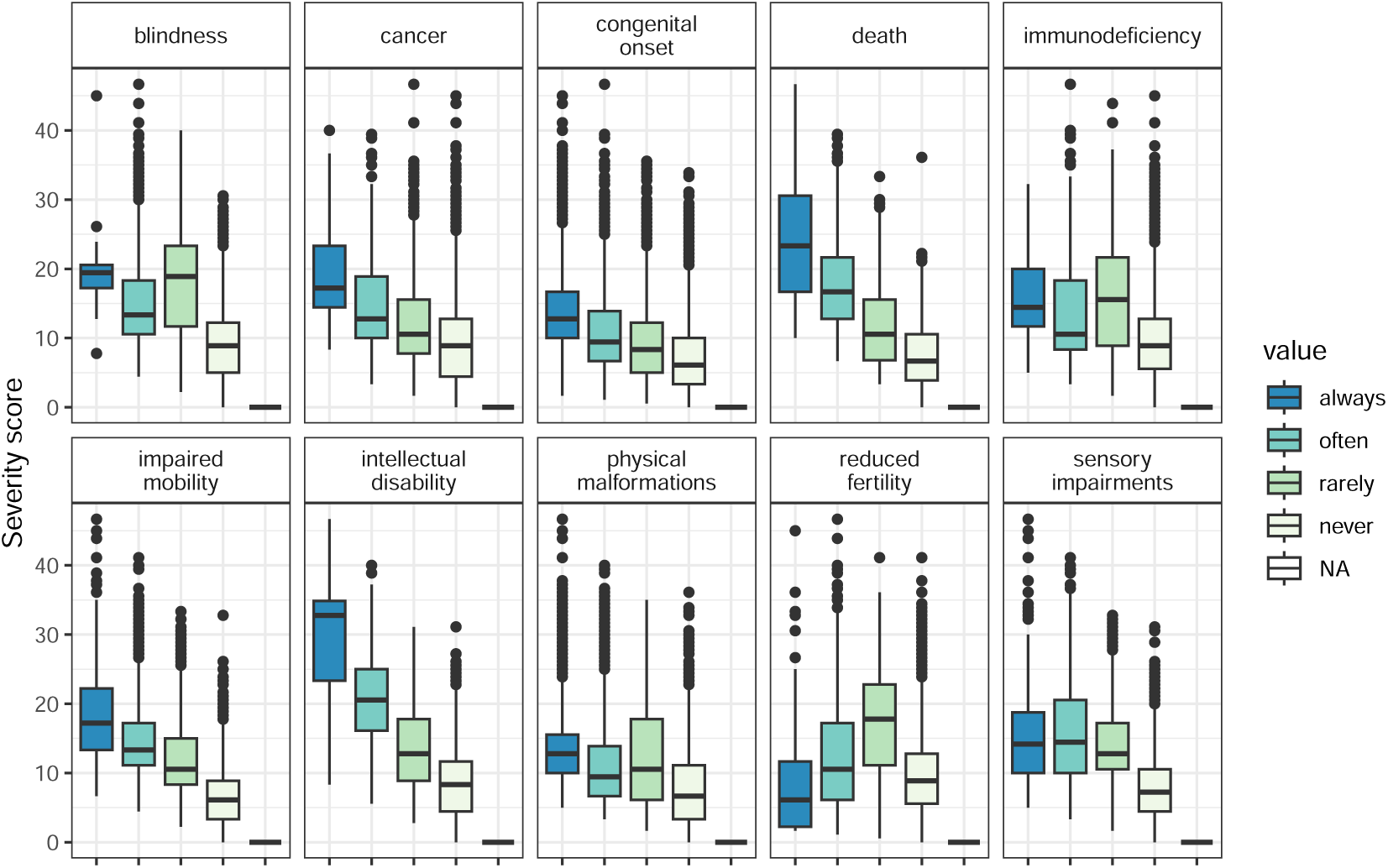
Boxplot showing the relationship between composite severity score (y-axis) and the frequency response categories within each clinical characteristic type.

**Figure 8:**
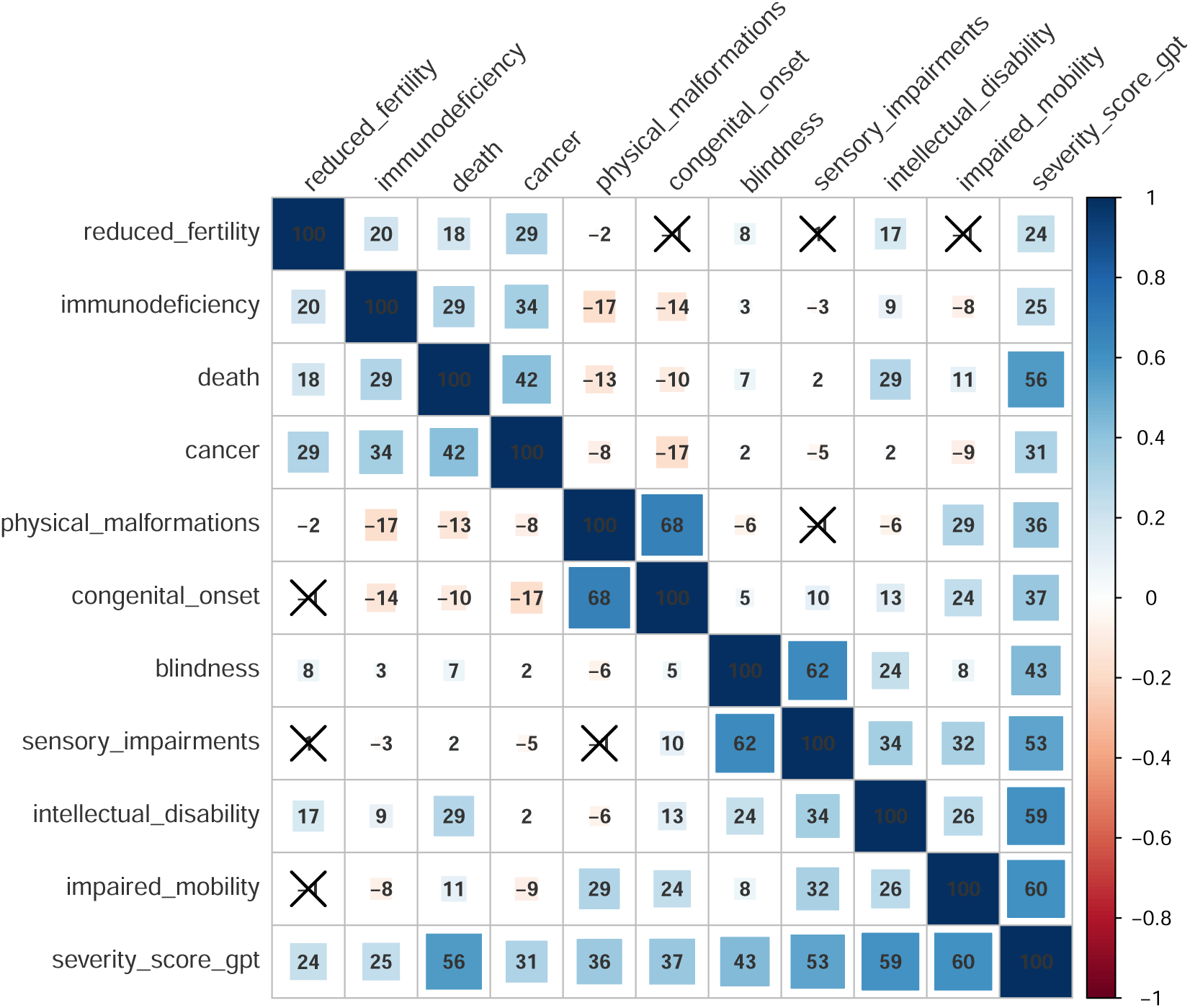
Pearson correlations between each individual clinical characteristic severity metric and the composite severity score (‘severity_score_gpt’).

**Figure 9:**
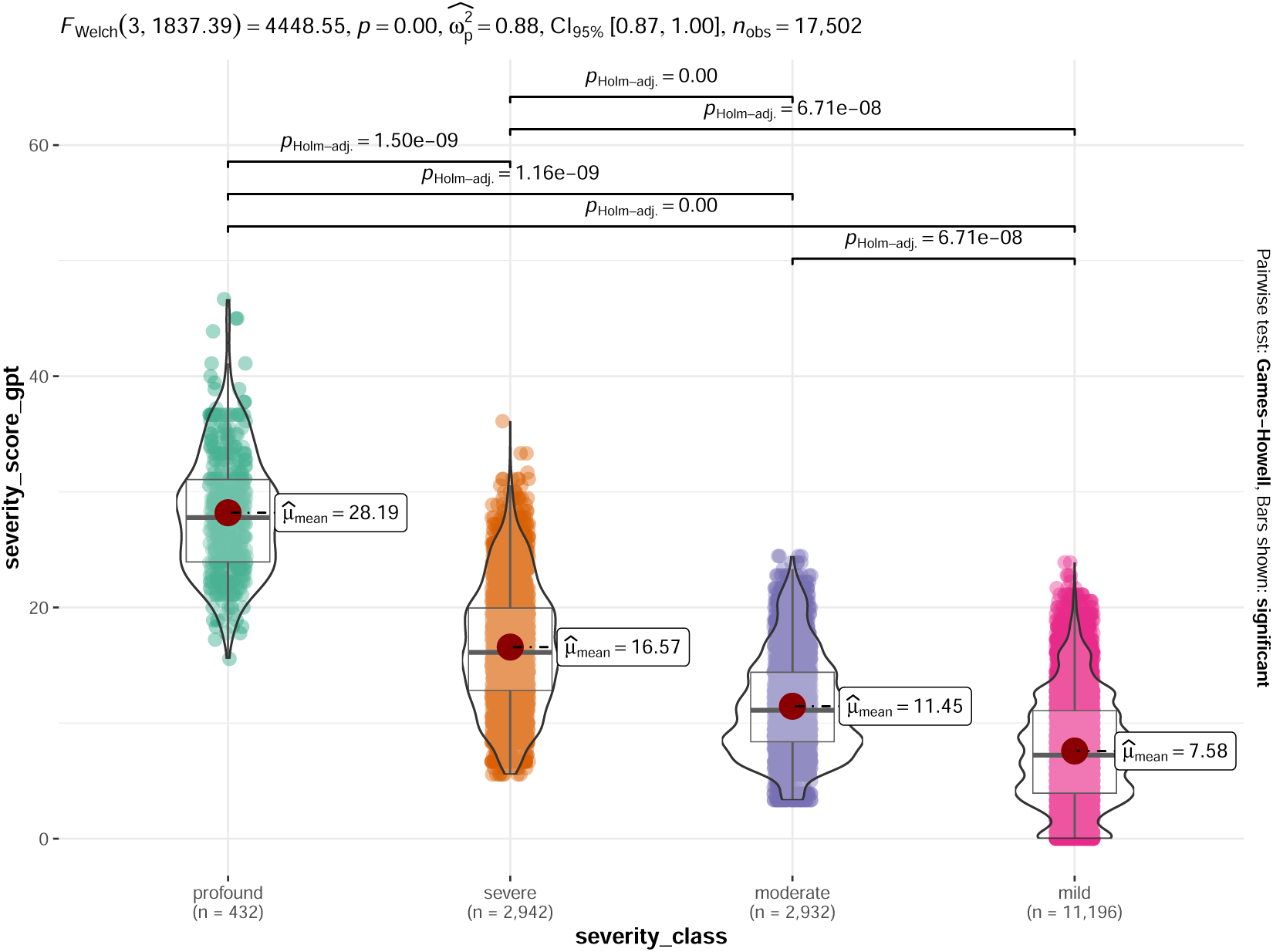
Distribution of the composite GPT-4 severity score introduced in this paper (y-axis) by an approximation of the severity class system introduced Lazarin et al. (2014) (x-axis). While these are different schemes for ranking phenotype severity, there is a strong correspondence between them (see summary statistics in subtitle). The sample size (number of phenotypes) is shown in parentheses along the x-axis.

**Table 3:** Table of GPT-4 clinical characterstic annotations for all Human Phenotype Ontology (HPO) phenotypes in Figure 3. For each phenotype, this includes the name of the phenotype (‘hpo_name’), the ID of the phenotype (‘hpo_id’), the frequency of each annotation (always, often, rarely, never), and the justification for each annotation (‘…_justification’). These results can also be downloaded programmatically using the R function HPOExplorer::gpt_annot_check.

**Top phenotype annotations table**

### 0.8.2 Supplementary Tables

**Table 4:** Severity nnotations generated for GPT-4 clinical characteristic annotations for the HPO phenotype ‘Fetal akinesia sequence’ (HP:000198).

| Clinical characteristic | Annotation | Justification |
| --- | --- | --- |
| Intellectual disability | always | Fetal akinesia sequence typically results in severe neurodevelopmental impairment, including intellectual disability. |
| Death | always | Fetal akinesia sequence is typically fatal in utero or shortly after birth. |
| Impaired mobility | always | Fetal akinesia sequence results in severe physical impairment, including impaired mobility. |
| Physical malformations | always | Fetal akinesia sequence is associated with multiple physical malformations. |
| Blindness | often | Visual impairment is common in surviving individuals with fetal akinesia sequence due to neurodevelopmental impairment. |
| Sensory impairments | always | Fetal akinesia sequence typically results in severe sensory impairment due to neurodevelopmental disruption. |
| Immunodeficiency | rarely | While not a direct feature, some individuals with fetal akinesia sequence may have associated immune abnormalities. |
| Cancer | never | Fetal akinesia sequence does not cause cancer. |
| Reduced fertility | often | Given the severe physical impairments associated with fetal akinesia sequence, fertility is likely to be reduced in surviving individuals. |
| Congenital onset | always | Fetal akinesia sequence is a congenital disorder. |

